# A longitudinal study of depressive symptom trajectories and risk factors in congestive heart failure

**DOI:** 10.1101/2024.09.16.24313783

**Authors:** Julia Gallucci, Justin Ng, Maria T. Secara, Brett D.M. Jones, Colin Hawco, M. Omair Husain, Nusrat Husain, Imran B. Chaudhry, Aristotle N. Voineskos, M. Ishrat Husain

## Abstract

**Background:** Depression is prevalent among patients with congestive heart failure (CHF) and is associated with increased mortality and healthcare utilization. However, most research has focused on high-income countries, leaving a gap in knowledge regarding the relationship between depression and CHF in low-to-middle-income countries (LMICs). This study aimed to delineate depressive symptom trajectories and identify potential risk factors for poor outcomes among CHF patients.

**Methods:** Longitudinal data from 783 patients with CHF from public hospitals in Karachi, Pakistan was analyzed. Depressive symptom severity was assessed using the Beck Depression Inventory (BDI). Baseline and 6-month follow-up BDI scores were clustered through Gaussian Mixture Modeling to identify distinct depressive symptom subgroups and extract trajectory labels. Further, a random forest algorithm was utilized to determine baseline demographic, clinical, and behavioral predictors for each trajectory.

**Results:** Four depressive symptom trajectories were identified: ‘good prognosis,’ ‘remitting course,’ ‘clinical worsening,’ and ‘persistent course.’ Risk factors associated with persistent depressive symptoms included lower quality of life and the New York Heart Association (NYHA) class 3 classification of CHF. Protective factors linked to a good prognosis included less disability and a non-NYHA class 3 classification of CHF.

**Conclusions:** By identifying key characteristics of patients at heightened risk of depression, clinicians can be aware of risk factors and better identify patients who may need greater monitoring and appropriate follow-up care.

**Clinical Perspective:** *What is new?:*

- To the best of our knowledge, this is the first study to use machine learning techniques to investigate depressive symptom trajectories in CHF patients from an LMIC.
- Four distinct depressive symptom trajectories were identified, ranging from good prognosis to persistent depressive symptoms.
- This study highlights protective and risk factors associated with these trajectories based on patients’ demographics and clinical presentations at baseline.

*What are the clinical implications?:*

- Personalized interventions based on identified protective factors for high-risk CHF patients could enhance both mental health and cardiovascular outcomes.
- Early detection and management of depression, particularly in patients with poor quality of life or advanced heart failure, may help reduce healthcare utilization and mortality.
- This study emphasizes the importance of routine depression screening in CHF patients, especially in LMICs, to enhance overall patient care and outcomes.

## 1. Introduction

Depression is common in congestive heart failure (CHF), with an estimated prevalence of 20-30% ^1,2^, making it one of the most frequent comorbid mental disorders in this patient population ^3^. Psychological health profoundly influences adherence to self-care practices ^4^, affecting both clinical outcomes and healthcare costs. Specifically, depression and depressive symptoms in CHF contributes to increased healthcare expenditures ^3,5^, higher rates of hospitalization ^6,7^, heightened heart failure symptoms ^6^ and is a risk factor for mortality ^3,7,8^. Depression and depressive symptoms in CHF are also linked to diminished quality of life ^3,9^, physical and social limitations ^6^, functional decline ^10^, worse New York Heart Association (NYHA) classification ^10^, and decreased compliance to treatment regimens ^3^. Despite the high prevalence and substantial relationship with adherence and outcomes, few studies have investigated depression and depressive symptoms in patients with CHF ^3^.

Much of our current understanding of biological, psychological, and social factors underlying depression and depressive symptoms in CHF is derived from studies conducted in high-income countries, where findings may not extend to other regions of the world given the differential and complex sociodemographic factors ^11^. A recent review highlighted social determinants such as socioeconomic status, race, and ethnicity as being influential in depression rates in CHF ^12^. Further, researchers have suggested an expected rise in CHF burden in low-to-middle-income countries (LMICs), where the etiology of CHF has been shown to vary by income level ^13^. The inclusion of diverse patient populations from LMICs can more comprehensively evaluate the relationship between depression and depressive symptoms in CHF, enhancing generalizability.

Previously, we prospectively investigated mortality, disability, and health-related quality of life in depressed patients with CHF from Pakistan, uncovering a high rate of depression and a correlation between the severity of depressive symptoms and increased mortality ^14^. Our earlier analyses were constrained by a binary classification of depression, which limited the ability to assess the heterogeneous impairments among patients and did not account for potential differential trajectories of depressive symptoms. Existing literature has identified depressive symptom trajectories in CHF patients from Western countries, consisting of various patterns of persistence, improvement, and worsening ^15–18^, however, to the best of our knowledge, these trajectories in CHF patients from LMICs are not known. Investigating these trajectories is crucial for enabling personalized care and early intervention, especially in regions with limited healthcare resources. Understanding these patterns can lead to more targeted treatment, efficient resource allocation, and culturally sensitive mental health strategies, ultimately improving both psychological and cardiac outcomes.

The present investigation leveraged a large longitudinal dataset of CHF patients from an urban setting in an LMIC (Karachi, Pakistan). Using machine learning techniques, we aimed to delineate depressive symptom trajectories among CHF patients and identify potential factors associated with these varying trajectories.

## 2. Methods

### 2.1 Data Source

Data was obtained from a 6-month cohort study on CHF and depression, detailed in our prior publication ^14^, involving 1009 hospital patients across the most populous city in Pakistan (Karachi) between January 4, 2010, and December 31, 2012, recently diagnosed with CHF. Participants completed assessments at baseline and after 6 months. Briefly, assessments included: social stress (Life Events Checklist; LEC), perceived support (Multidimensional Scale of Perceived Social Support; MSPSS), quality of life (Euro Quality of Life Visual Analog Scale; EQVAS), disability (Brief Disability Questionnaire; BDQ), health factors such as depressive symptom severity (Beck Depression Inventory; BDI), etiology of CHF, presence of left ventricular failure (LVF), CHF class (NYHA classification), smoking status, ejection fraction, and comorbidities (i.e. diabetes, chronic obstructive pulmonary disease; COPD, stroke, myocardial infarction, bypass surgery, and renal disease).

### 2.2 Statistical Analysis

Our statistical methods workflow is outlined in **Figure 1** and involved two main aims: 1) to identify depressive symptom trajectories and 2) to identify factors contributing to these depressive symptom trajectories.

**Figure 1:**
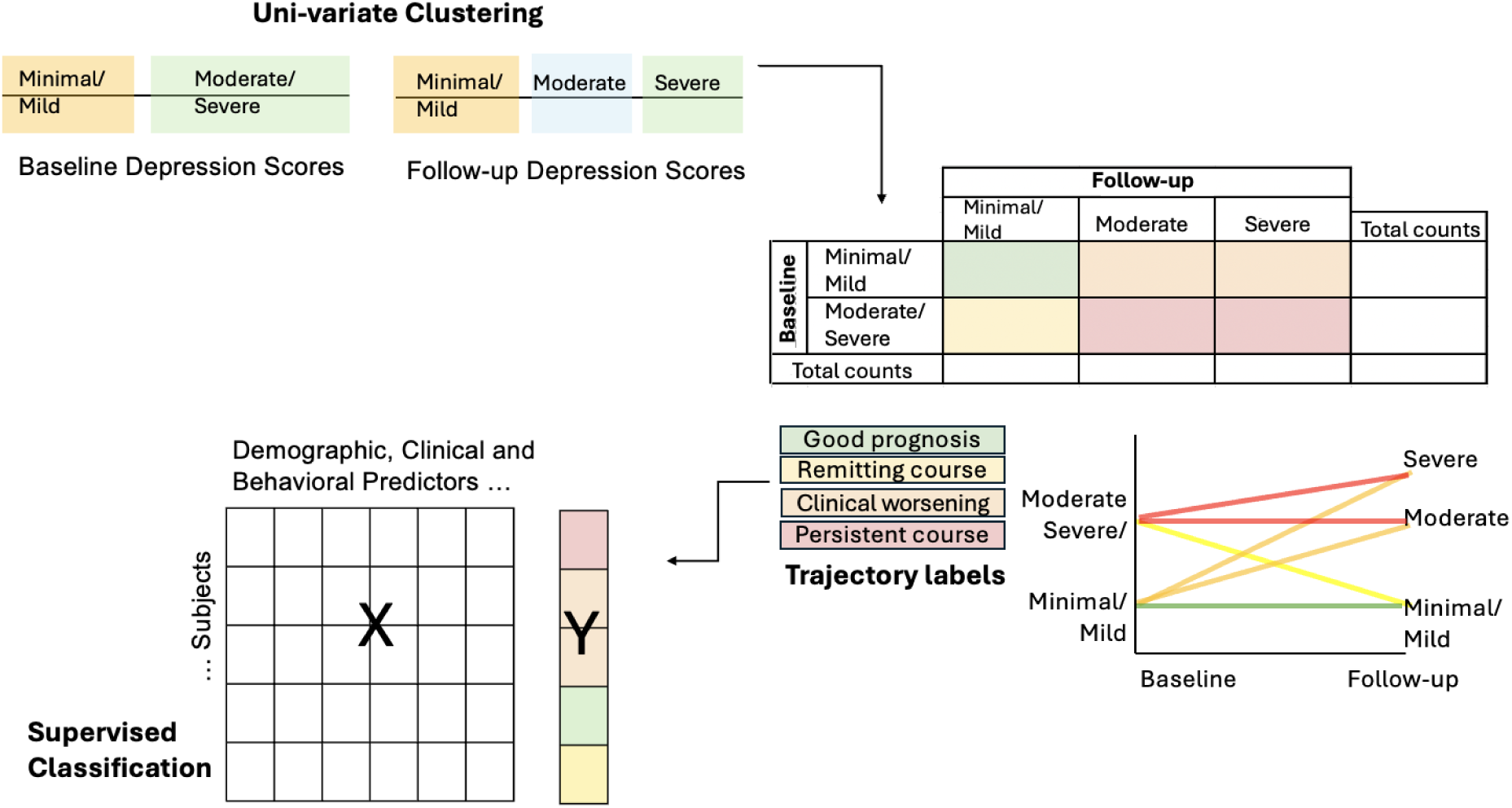
Schematic of Analysis. Using univariate clustering, the baseline and follow-up depressive symptom severity scores (BDI) were analyzed separately to identify distinct clusters. From this, individual trajectory labels were assigned to participants based on these clusters, and a supervised classifier was applied to uncover demographic, clinical, and behavioral factors predictive of each trajectory label.

#### 2.2.1 Aim 1: Identify depressive symptom trajectories

To identify depressive symptom trajectories, a Gaussian Mixture Model (GMM) clustering approach was employed separately on baseline and follow-up depressive symptom severity scores (BDI). Silhouette analysis was conducted to determine the optimal number of clusters. Silhouette scores measure the similarity of an object to its own cluster compared to others, in which higher scores indicate better cluster separation. Trajectory labels were extracted based on the cluster assignment.

#### 2.2.2 Aim 2: Identify factors contributing to depressive symptom trajectories

A random forest classifier was used to identify baseline features contributing to depressive symptom trajectories. A one-vs-rest approach was implemented, employing separate binary classifiers to distinguish each class from the others. This method is particularly effective for imbalanced classes and provides insights into the specific characteristics of each trajectory. All available baseline clinical and demographic variables were included in the model for a data-driven approach to determining relevant factors. Baseline depressive symptom severity (BDI) was omitted from our model to prevent data leakage, as clustering was based on this variable. Random forest was leveraged due to its employment of multiple decision trees, which effectively minimize uncertainty in prediction labels and reduce variance (i.e., sensitivity to idiosyncrasies in the training data). This ensemble method uses bootstrapping and random feature selection to enhance the independence of individual trees and facilitates the quantification of feature importance.

#### 2.2.3 Hyperparameter Optimization

To improve model performance, we used a nested cross-validation approach with 5 inner folds. The inner loop of nested cross-validation performs hyperparameter tuning by further splitting the training data into inner training and validation folds. We used the Tree-Structured Parzen Estimator (TPE) algorithm, which randomly initialized sets of hyperparameters to evaluate, then adaptively selected subsequent sets based on feedback to improve model performance. We conducted 500 evaluations, optimizing hyperparameters for the F1 score, the harmonic mean of precision, and recall, where precision is the proportion of correctly predicted positive labels among all predicted positives and recall is the proportion of correctly predicted positive labels among all actual positives. The number of trees was set to 500 to balance performance and computational cost. Key hyperparameters optimized included the splitting criterion (Gini impurity or Shannon information gain), class weight for imbalance, maximum tree depth, maximum features per split, and minimum samples per leaf and split.

#### 2.2.4 Performance Metrics

We employed multiple metrics to comprehensively evaluate classifier performance. Besides the F1 score, confusion matrices were normalized to show the percentage of true labels classified as each predicted label. Accuracy was calculated as the ratio of correctly identified labels to the total number of labels. Additionally, we assessed the area under the precision-recall curve (AUC-PR) and the area under the receiver operating characteristic curve (AUC-ROC) to measure the trade-offs between precision and recall, and true positive rate and false positive rate, respectively. The outer loop of nested cross-validation estimates model performance on unseen data, preventing overfitting during hyperparameter tuning and reducing the risk of biased performance estimates compared to a single random train-test split. Each model’s performance on new data was estimated using 10-folds with 5 repetitions. This approach involved multiple splits of the data into training and testing folds. By averaging performance metrics across folds, this provided an unbiased assessment of the model’s overall effectiveness.

#### 2.2.5 Feature Importance and Directionality

Feature importance was evaluated by quantifying each feature’s capacity to reduce impurity, thereby decreasing label uncertainty during decision tree construction. The reduction in impurity attributed to each feature was averaged across all trees in the random forest to determine its overall importance.

Although feature importance quantifies the extent to which each feature contributes to the prediction, it does not convey the direction of this influence. Shapley Additive exPlanations (SHAP) values were used to address this ^19^. SHAP values provide a detailed account of each feature’s contribution to a prediction. Each feature is assigned a SHAP value for every prediction, reflecting both the magnitude and direction of its impact. A higher SHAP value indicates a greater role of the feature in influencing the prediction. A positive SHAP value signifies that an increase in the feature’s value drives the prediction toward the current class, whereas a negative SHAP value suggests that an increase in the feature’s value moves the prediction away from the current class.

#### 2.2.6 Statistical Testing

To assess if the models predicted depressive symptom trajectories better than statistical chance, we performed non-parametric permutation testing on the performance metric for each model. Specifically, we conducted statistical testing on the F1 score to maintain consistency with hyperparameter optimization. To determine whether each feature contributed more to the prediction than statistical chance, we performed nonparametric permutation testing on the feature importance for each model. This involved randomly shuffling class labels outside the cross-validation framework and repeating cross-validation with the same previously optimized hyperparameters. We repeated this 1000 times, generating permuted null distributions to compare the true F1 and feature importance scores. We conducted one-sided tests to determine the number of permuted F1 and feature importance scores that exceeded the true scores at a significance level of α = 0.05. To correct for multiple comparisons, we separately conducted Benjamini-Hochberg FDR-correction ^20^ across the performance metric tests (4 x 1 comparisons) and the feature importance tests (4 x 32 comparisons) ^21^.

### 2.3 Code Sharing

All code used for statistical analyses and generating figures can be found at https://github.com/juliagallucci/CHF_Depressive_Symptoms.

## 3. Results

### 3.1 Participant Characteristics

A total of 783 individuals diagnosed with CHF were included in the analysis, all with complete data, including baseline and follow-up BDI scores, as well as all baseline demographic, behavioral, and clinical variables. Participant demographics and clinical scores are detailed in **Table 1**.

**Table 1:**
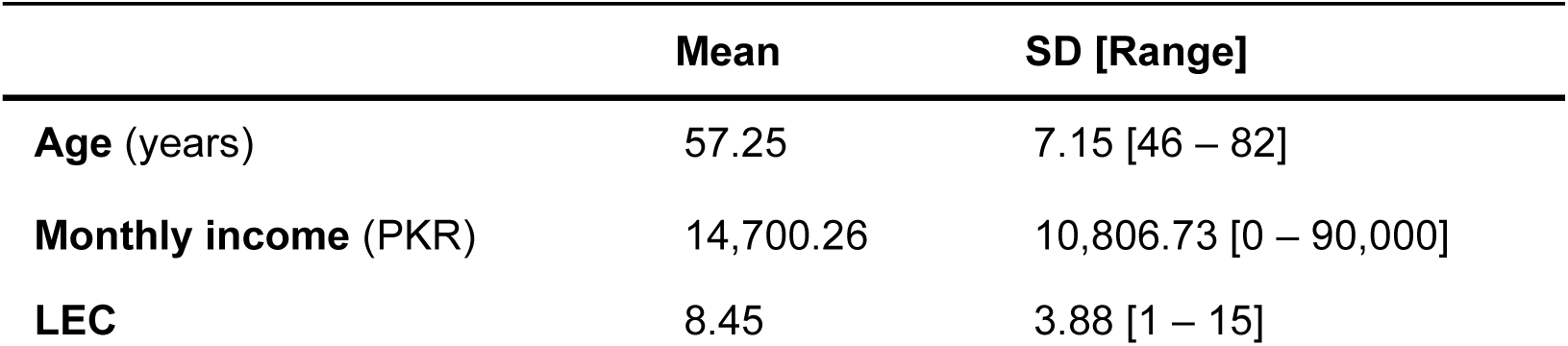
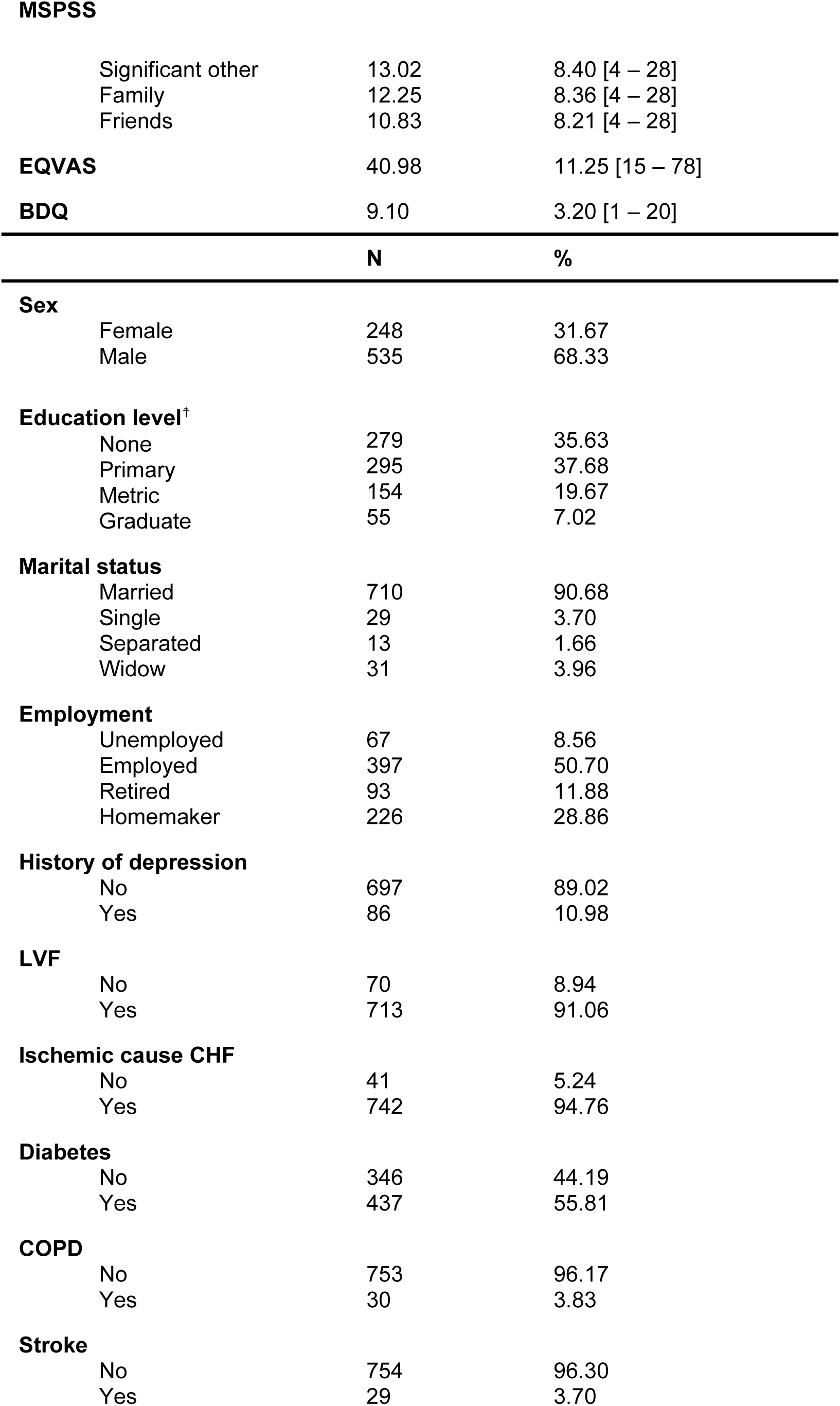
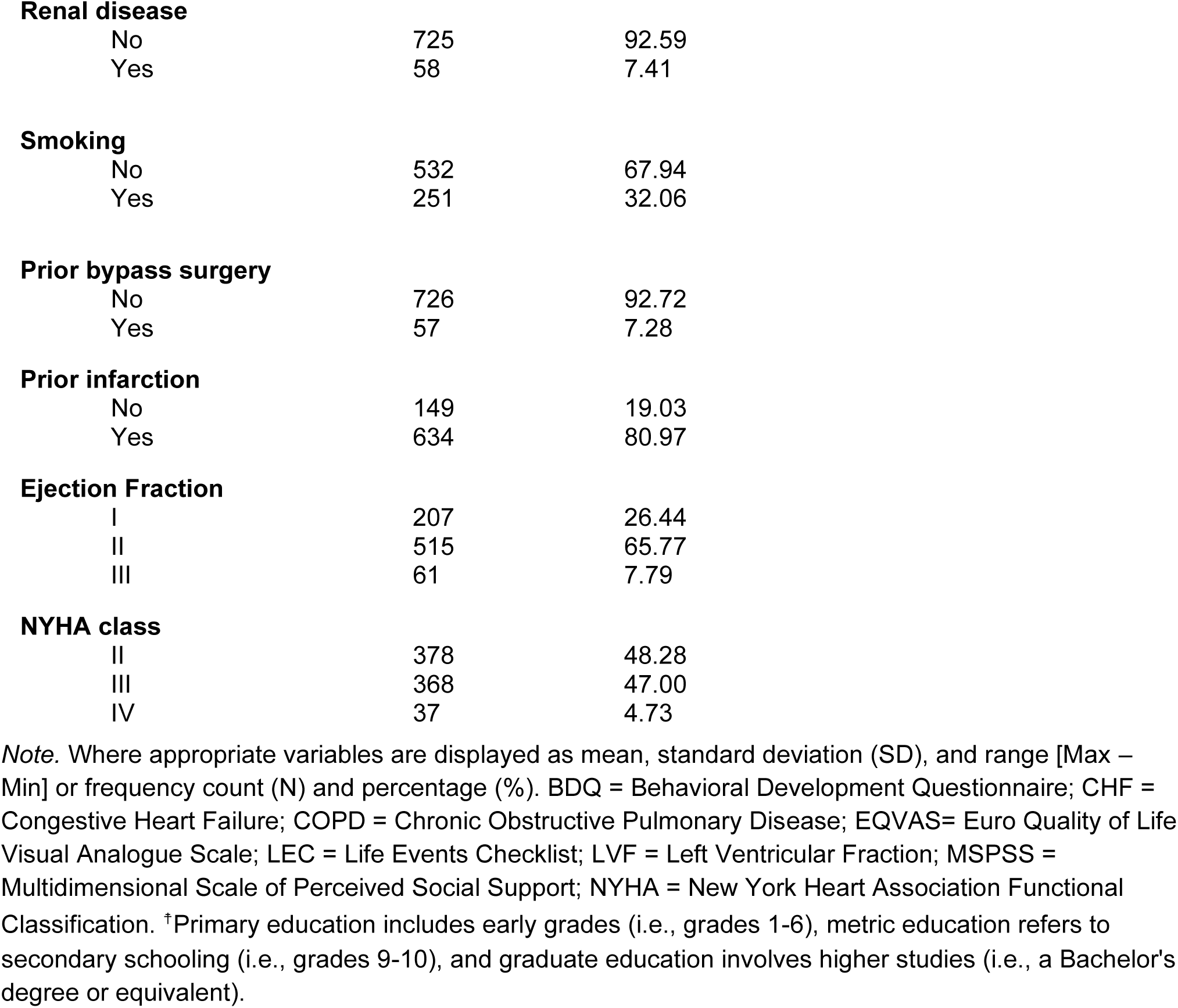
Participant’s Demographics and Clinical Score (n = 783)

### 3.2 Identifying Depressive Symptom Trajectories

#### 3.2.1 Clustering at Baseline and Follow-up

The GMM algorithm determined two distinct patient clusters based on depressive symptom severity (BDI) at baseline: ‘minimal/mild’ (30.65%), and ‘moderate/severe’ (69.35%) depressive symptom severity, based on the highest silhouette score (0.72) (see **Figure 2A**). At follow-up, three distinct patient clusters emerged: ‘minimal/mild’ (42.02%), ‘moderate’ (38.06%), and ‘severe’ (19.92%) depressive symptom severity, based on the highest silhouette score (0.68) (see **Figure 2B**).

**Figure 2.**
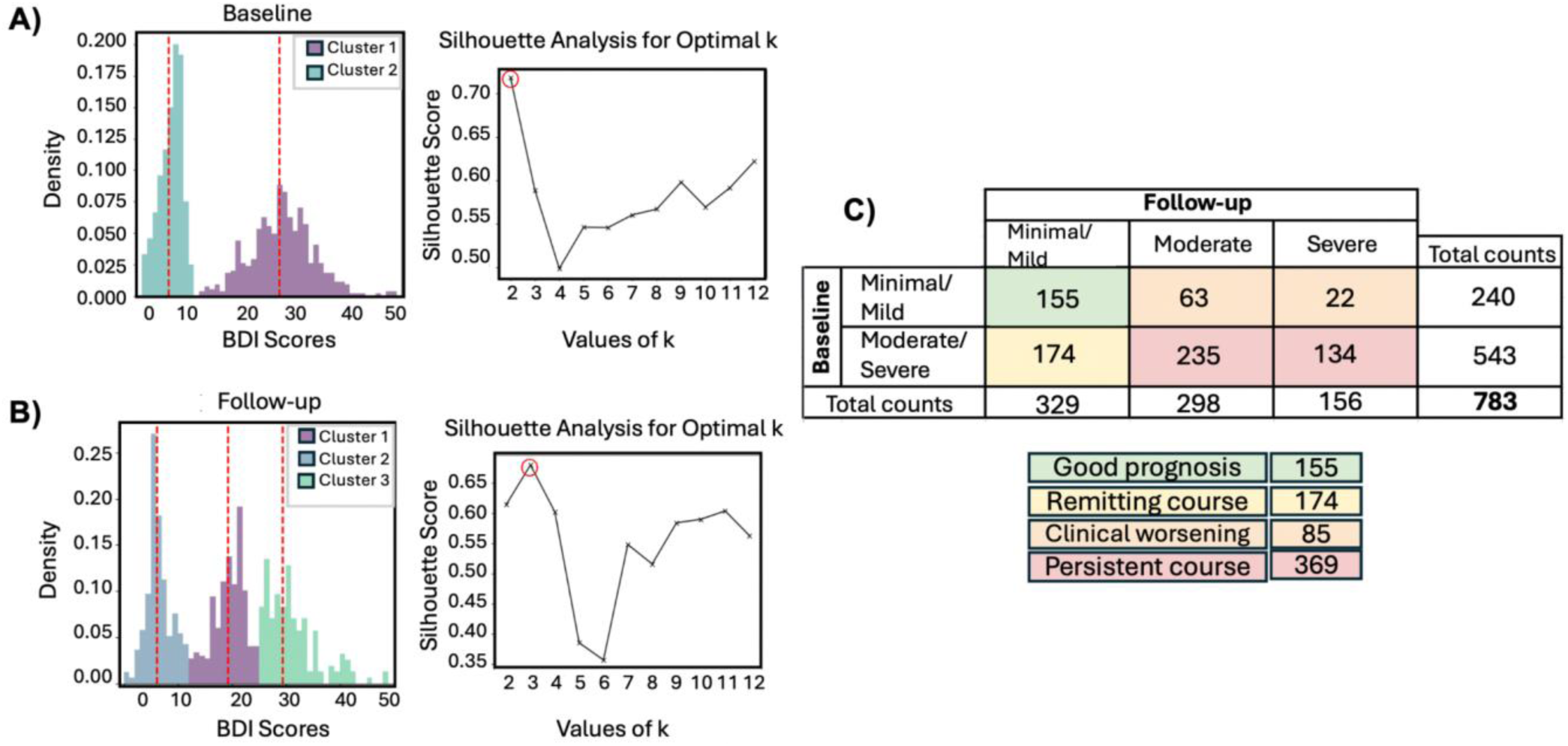
Depressive symptom trajectories of patients with CHF. GMM clustering and silhouette analysis were used to determine the optimal number of clusters at baseline **(A)** and at 6-month follow-up **(B)**. **C)** From these analyses, six-month trajectory labels of depressive symptoms were derived.

#### 3.2.2 Applying Trajectory Labels

Four trajectories were identified: ‘good prognosis’ (i.e. minimal/mild at baseline and follow-up; n = 155), ‘remitting course’ (i.e. moderate/severe at baseline and minimal/mild at follow-up; n = 174), ‘clinical worsening’ (i.e. minimal/mild at baseline and either moderate or severe at follow-up; n = 85), and ‘persistent course’ (i.e. moderate/severe at baseline and follow-up; n = 369) (see **Figure 2C**). Each trajectory’s demographic and clinical characteristics are presented in Supplemental Table S1.

### 3.3 Factors Contributing to Depressive Symptom Trajectories

The initial analysis involved implementing the random forest classifier with default hyperparameters. While the model accurately predicted ‘good prognosis’ and ‘persistent course,’ it frequently misclassified ‘clinical worsening’ and ‘remitting course’ (Supplemental Figure S1). To address this, we optimized hyperparameters to improve the F1 score, which was particularly sensitive to these misclassifications. Given the relatively good classifier performance (all p<0.0099), with the confusion matrix showing the majority of true labels correctly classified and few misclassifications (**Figure 3**), the most critical features at baseline for each outcome using the F1-optimized models were investigated (**Figure 4**).

**Figure 3.**
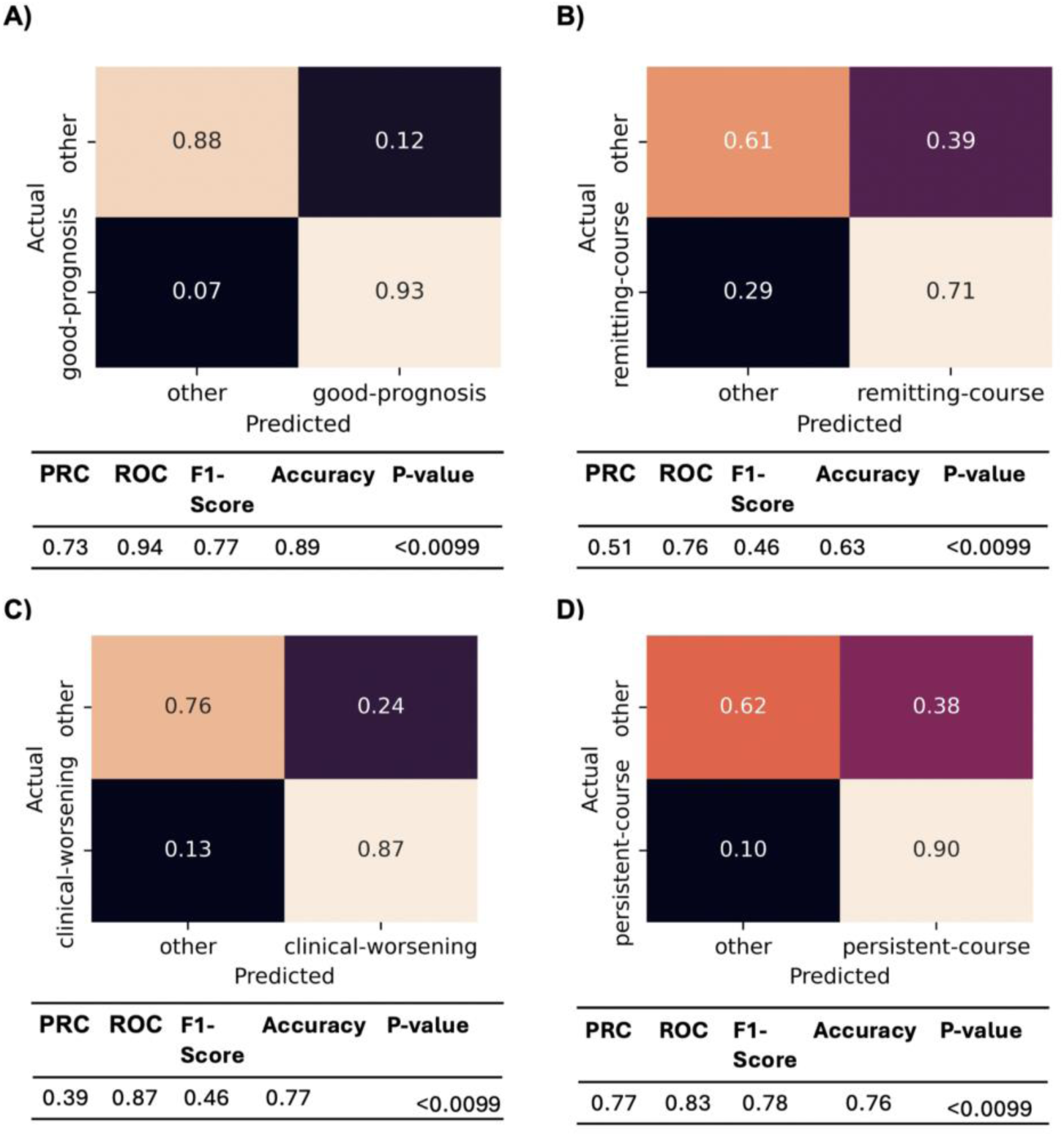
Confusion Matrices and Performance Metrics of One-vs-Rest Binary Classifiers Optimized for F1. ‘Good prognosis’ **(A)**, ‘remitting course’ **(B)**, ‘clinical worsening’ **(C)**, and ‘persistent course’ **(D)** labels.

**Figure 4.**
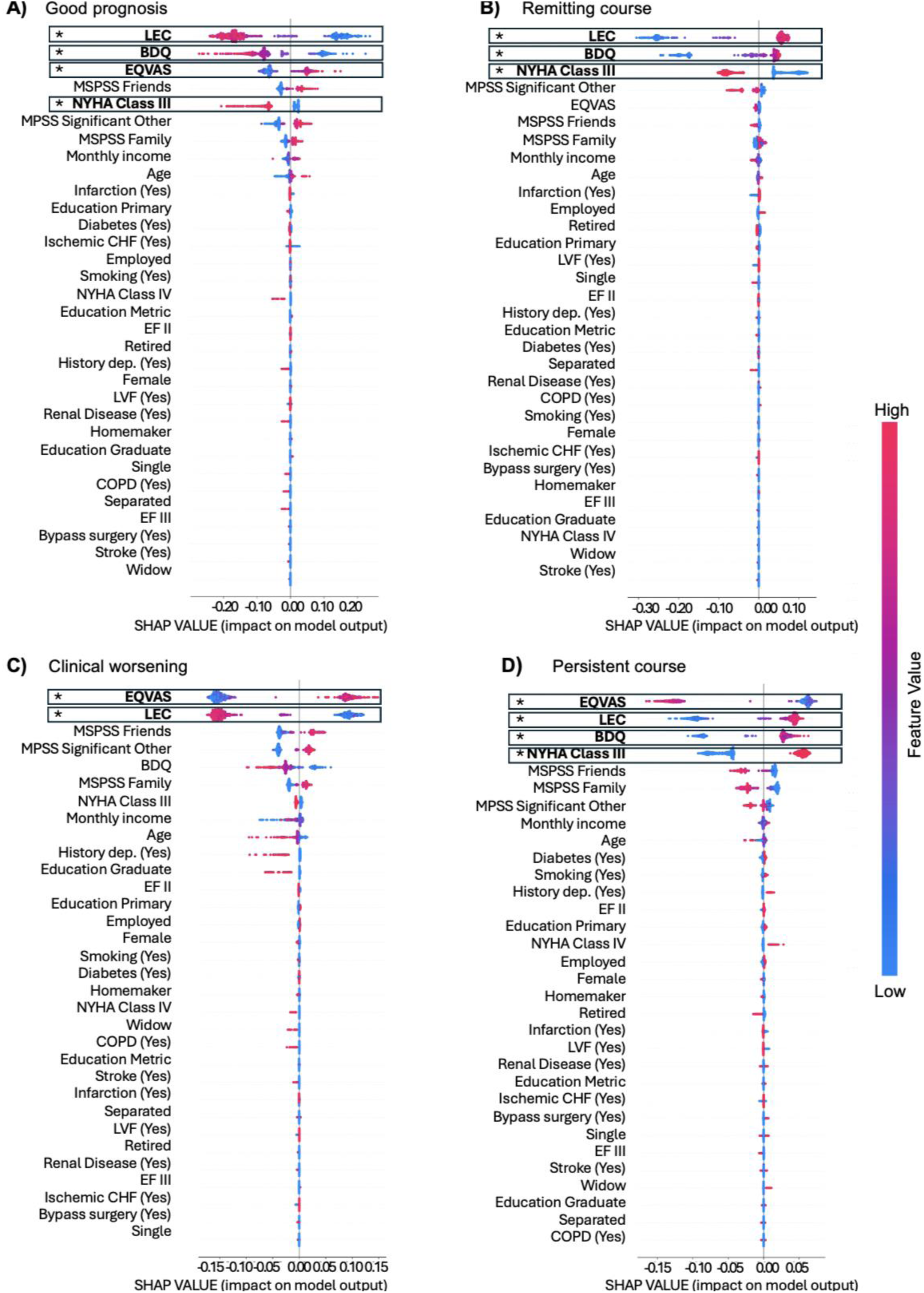
SHAP values for one-vs-rest random forest models. SHAP (SHapley Additive exPlanations) values for predictors of ‘good prognosis’ (**A**), ‘remitting course’ (**B**), ‘clinical worsening’ (**C**), and ‘persistent course’ (**D**). Positive SHAP values indicate a positive contribution to the prediction, while negative values indicate a negative contribution. Predictors are ordered based on impurity-based feature importance, with significance indicated (* pFDR < 0.05).

Key features for a ‘good prognosis’ included a positive association with quality of life (feature importance 0.15; pFDR = 0.014) and negative associations with social stress (feature importance 0.39; pFDR = 0.014), disability (feature importance 0.18; pFDR = 0.014), and NYHA class 3 (feature importance 0.06; pFDR = 0.049) (**Figure 4A**). Key features for a ‘remitting course’ included positive associations with social stress (feature importance 0.40; pFDR = 0.014) and disability (feature importance 0.28; pFDR = 0.014) and a negative association with NYHA class 3 (feature importance 0.13; pFDR = 0.026) (**Figure 4B**). Key features of ‘clinical worsening’ included a positive association with quality of life (feature importance 0.32; pFDR = 0.014) and a negative association with social stress (feature importance 0.31; pFDR = 0.014) (**Figure 4C**).

Lastly, key features of a ‘persistent course’ included positive associations with social stress (feature importance 0.20; pFDR = 0.014), disability (feature importance 0.17; pFDR = 0.046), and NYHA class 3 (feature importance 0.12; pFDR = 0.049), and a negative association with quality of life (feature importance 0.26; pFDR = 0.014) (**Figure 4D**).

To better interpret these factors, trajectories were grouped post-hoc based on similar baseline depressive symptom severity scores (**Figure 5**). For trajectories with minimal baseline depressive symptom severity (‘good prognosis’ and ‘clinical worsening’), quality of life had a positive association, and social stress had a negative association. Uniquely, maintaining minimal depressive symptom severity over time (‘good prognosis’) was negatively associated with disability and NYHA class 3. In contrast, trajectories with moderate/severe baseline depressive symptom severity (‘persistent course’ and ‘remitting course’) had positive associations with both disability and social stress. Uniquely, a poor prognosis over time (‘persistent course’) was negatively associated with quality of life and positively associated with NYHA class 3, while symptom improvement over time (‘remitting course’) was negatively associated with NYHA class 3.

**Figure 5.**
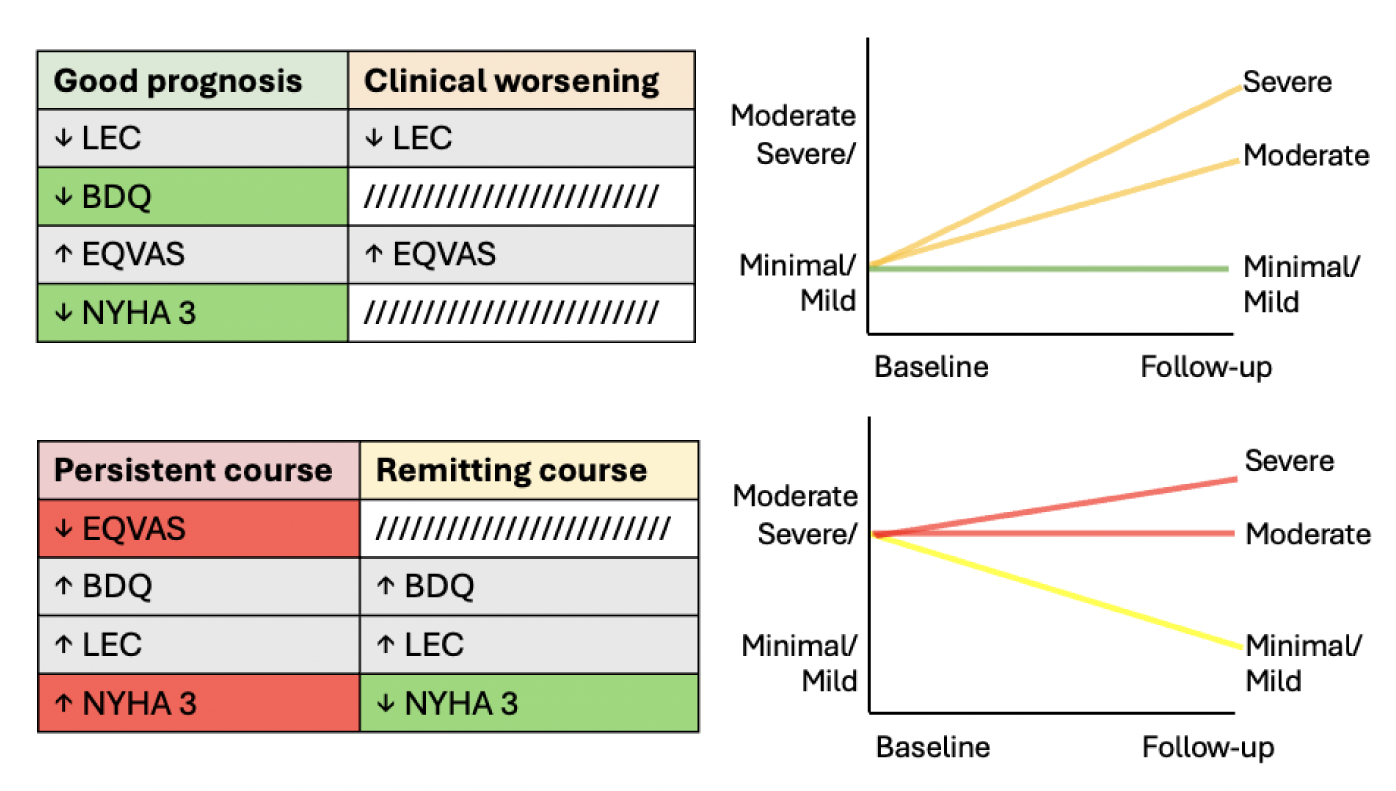
Significant predictors and their associations with trajectories, grouped based on shared baseline depressive symptom severity scores. Risk factors (red) are uniquely linked to poorer outcomes, protective factors (green) are uniquely linked to better outcomes, and common factors (grey) are associated with both.

## 4. Discussion

In the present investigation, we leveraged a longitudinal dataset of CHF patients from an urban setting in an LMIC that was also assessed for depressive symptoms. We aimed to uncover patterns of depressive symptom severity over time and explore related baseline demographics and clinical factors. We identified four distinct depressive symptom trajectories: ‘good prognosis,’ ‘remitting course,’ ‘clinical worsening,’ and ‘persistent course.’ Key contributors across these trajectories included quality of life, social stress, disability, and CHF classification. Specifically, minimal baseline depressive symptom severity was linked to better quality of life and lower social stress, while lower disability levels and a non-NYHA class 3 classification were associated with good longitudinal outcomes. Conversely, moderate/severe baseline depressive symptom severity was linked to higher disability and social stress, while lower quality of life and NYHA class 3 classification were associated with more persistent poorer outcomes.

Our study identified four distinct depressive symptom trajectories in this sample. Approximately half of those presenting with depressive symptoms at baseline persist at follow-up, while around 40% either maintain minimal symptoms or exhibit clinical improvement. A small percentage show worsening of depressive symptoms after six months. These results are consistent with previous research on depressive symptom trajectories in Western countries ^15–18^, highlighting that a significant portion of CHF patients experience persistent depressive symptoms and among those who do show changes, the majority tend to improve rather than decline, irrespective of setting.

Poorer longitudinal outcomes were linked to a lower quality of life and a higher CHF functional classification (NYHA class 3), indicating moderately severe cardiovascular disease with physical activity limitations. These findings are consistent with studies linking higher NYHA class to increased psychological distress, such as anxiety ^22^, depression ^23,24^, and overall reduced quality of life ^25,26^. Given the subjective nature of NYHA, which is based on patients’ experience of limitations in daily activities due to cardiac function ^22,26^, persistent depressive symptoms may distort illness perception, leading to an overemphasis on functional limitations. This interplay may worsen the perception of heart failure, leading CHF patients with persistent depressive symptoms to report greater limitations and poorer functioning, which in turn can perpetuate their depressive symptoms. Relatedly, it has been shown that the perception of symptom burden significantly affects mood-related symptoms (e.g., anxiety and depression) in patients with CHF ^27,28^.

Lower disability and non-severe CHF functional classification (i.e., non-NYHA class 3) were identified as protective factors against poorer longitudinal outcomes. Trajectories characterized by worsening or persistent clinical symptoms lack these protective factors, which may lead to less favorable outcomes. Increased disability is associated with decreased quality of life and is an independent predictor of mortality ^29,30^. Additionally, disability directly correlates with NYHA functional classification and is independently linked to cognitive dysfunction in CHF patients ^31^. These associations can impair patients’ ability to adhere to treatments, manage self-care, and make informed health-related decisions ^32^, potentially contributing to a cycle of worsening physical and psychological health outcomes. Trajectories with more favorable outcomes (i.e., ‘good prognosis’ and ‘remitting course’) show a negative association with NYHA class 3 and disability, particularly in the ‘good prognosis’ group. These findings suggest that the level of disability, affecting physical and cognitive function as well as emotional well-being, may directly influence longitudinal outcomes and mediate the transition between favorable and unfavorable trajectories. Interventions such as resistance training and personalized exercise programs could help reduce physical disability ^33,34^ and improve cognitive functioning ^35^. Exercise has also shown efficacy in reducing depression ^36^, which may improve patient trajectories for individuals with CHF experiencing depressive symptoms.

This study has some limitations. Our analyses lacked an independent dataset to evaluate the generalizability of results. However, cross-validation and permutation procedures were employed to mitigate the risk of unstable findings. Additionally, this study was conducted as a secondary analysis and therefore was not originally designed prospectively to address the proposed aims. For instance, our data collection was limited to baseline and 6-month follow-up assessments. Serial assessments of depressive symptoms spanning this period would offer a more nuanced understanding of individuals’ true depressive symptom trajectories. The association between more severe NYHA classifications and depressive symptoms could indicate that severe CHF causes depression in patients with significant functional impairment, or that depressive symptoms worsens heart failure symptoms and functional impairment. The study cannot establish the causality of this complex bidirectional relationship and requires further analysis to explore whether this association could help explain the link between depression and increased mortality in CHF patients.^37^. Lastly, while we relied on the BDI, a widely accepted measure of depressive symptom severity in adults ^38^, it lacks the diagnostic capabilities of gold standard instruments like the Diagnostic and Statistical Manual of Mental Disorders (DSM) or Mini International Neuropsychiatric Interview (MINI). This limitation, coupled with the absence of assessments for other psychopathologies such as anxiety or substance use disorders, restricts our ability to fully attribute observed outcomes to depression alone. Including a broader range of assessments could have provided a more comprehensive understanding of how these factors interact with physical health outcomes.

The present study underscores the heterogeneous nature of depressive symptom trajectories among CHF patients in LMICs, revealing a complex interplay of contributing risk and protective factors. These findings are strengthened by using a data-driven clustering approach, which led to a set of distinct trajectory clusters, rather than a binary classification of depression status. This method provided a more nuanced understanding of depressive symptom trajectories by identifying natural groupings within the data, leading to more accurate and personalized insights into patient outcomes that a depressed vs. non-depressed dichotomy might overlook. Additionally, the use of a random forest classifier, a method robust to overfitting and capable of handling large imbalanced datasets, proved instrumental in uncovering meaningful predictors for each trajectory. Such risk factors must be carefully considered to enhance understanding of patients at heightened risk of adverse outcomes, facilitating early identification and expediting initiation of appropriate interventions to maximize benefits. Furthermore, our findings may inform clinical trials aimed at preventing depression development and effectively managing diagnosed cases, ultimately improving patient outcomes through tailored interventions. For example, given the relationship between physical disability and depression, patients at high risk might be enrolled in more intensive rehabilitation and exercise programs. This is particularly important given the significant impact of depression on mortality in CHF patients ^3,7,8^, notwithstanding the additional suffering associated with comorbid conditions ^39^.

## Data Availability

The data referred to in this manuscript is not publicly available due to privacy restrictions. However, the code used for analysis is provided to ensure reproducibility and transparency.

https://github.com/juliagallucci/CHF_Depressive_Symptoms

## Acknowledgments

The data utilized in this work was collected by MIH. The analysis was supported by Rahul Krishnan.

